# De-identifying Spanish medical texts - Named Entity Recognition applied to radiology reports

**DOI:** 10.1101/2020.04.09.20058958

**Authors:** Irene Pérez-Díez, Raúl Pérez-Moraga, Adolfo López-Cerdán, Marisa Caparrós Redondo, Jose-Maria Salinas-Serrano, María de la Iglesia-Vayá

## Abstract

Medical texts such as radiology reports or electronic health records are a powerful source of data for researchers. Anonymization methods must be developed to de-identify documents containing personal information from both patients and medical staff. Although currently there are several anonymization strategies for the English language, they are also language-dependent. Here, we introduce a named entity recognition strategy for Spanish medical texts, translatable to other languages. We tested 4 neural networks on our radiology reports dataset, achieving a recall of 97.18% of the identifying entities. Alongside, we developed a randomization algorithm to substitute the detected entities with new ones from the same category, making it virtually impossible to differentiate real data from synthetic data. The three best architectures were tested with the MEDDOCAN challenge dataset of electronic health records as an external test, achieving a recall of 69.18%. The strategy proposed, combining named entity recognition tasks with randomization of entities, is suitable for Spanish radiology reports. It does not require a big training corpus, thus it can be easily extended to other languages and medical texts, such as electronic health records.

## Background

Medical imaging is widely used in clinical practice for the diagnosis and treatment of several diseases, such as alzheimer, cancer or pneumothorax. Data from radiology reports, electronic health records and other medical texts such as clinical trial protocols are being used for research purposes (1, 2). Health care institutions, researchers and patients can greatly benefit from these datasets. However, these records and reports contain patient notes known as personal data that can challenge patient confidentiality and privacy, as provided for in the European Regulation on the protection of personal data (3). All words that could identify a patient must be removed or de-identified before data analysts start their research or even more before the dataset is published.

From a legal point of view, Regulation (EU) 2016/67 on the protection of natural persons and with regard to the processing of personal data and on the free movement of such data (3) provides the regulatory framework in the European Union. Although its application is mandatory to all its member states, its concrete implementation varies depending on each of them. In Spain, the Organic Law 3/2018 (4) establishes the legal framework for data protection in biomedical research. Reuse of personal data for medical research needs to be approved by an ethics committee, and data must be at least pseudonymized before the researchers get access to it. Legal issues regarding data privacy are not the only source of concern. Direct consequences to patients are also a very important factor to be carefully considered. It is crucial to protect the private health details of a patient from any third party’s access, and avoid exposing identifiable personal data such as identifier numbers or addresses. De-identification is therefore essential to ensure patient privacy and comply with legal requirements.

From a data management point of view, the de-identification methodology needs to be precise and recallable. Precision is needed to minimize the data loss of the de-identification process and to preserve the semantic meaning of the radiology report; recall allows getting the best de-identification possible and avoiding losing any identifiable information (5).

Even though several de-identification or anonymization methodologies have been proposed in English, legislation differs on a national level worldwide and language-specific problems can arise, hence a different method for each language must be developed. These difficulties extend to any Natural Language Processing (NLP) implementation. In the biomedical field, NLP has been applied successfully in English, including for de-identification purposes (6), but many of these strategies rely on language-specific resources and are not extensible to other languages (7). Apart from the English language, this problem has been assessed in French, where different strategies from machine learning to the use of dictionaries and lists have been proposed, along with protocols for corpus development (8, 9). In other languages such as German, Swedish, Dutch or Chinese some strategies and methodologies have also been proposed (5, 10–13), but there have been so far rather limited attempts in automatic de-identification for Spanish medical texts (14, 15), including the MEDDOCAN task (16).

Most of the works around text de-identification are based on pattern matching or machine learning, or even a combination of both. Whereas pattern matching does not account for the context of a word and is unaware of typographical errors, machine learning techniques require a large corpus of annotated text (17). Since our radiology reports were mostly free text with sensible data outside headers, we opted for annotating our own corpus and developing a Named Entity Recognition (NER) based de-identification method.

## Methods

### Named Entities

Given that there is no specific guidance in the Spanish legal system on what information has to be removed to deidentify medical texts, we decided to search in our reports for the Protected Health Information (PHI) categories defined by the Health Insurance Portability and Accountability Act (HIPAA) in the United States of America (18). After manual inspection of the data and considering the scope of this work, we performed a sub-selection of PHI categories and finally grouped them in 6 Named Entities (NEs) as shown in Table 1. Some NEs included other information that should be protected to preserve the privacy of patients or doctors but was not included in PHI categories, such as digital signatures or healthcare centres.

**Table 1.** Named Entities selected for this task and their associated Protected Health Information categories

| NEs | Description | PHIs |
| --- | --- | --- |
| CAB | Section headers | - |
| NAME | Names and surnames (patient and others) | Names |
| DIR | Full addresses, including streets, numbers and zip codes | Geographic data |
| LOC | Cities, inside and outside addresses | Geographic data |
| NUM | Numbers or alphanumeric strings that might identify someone, including digital signatures, patient numbers, medical numbers, medical license numbers and others | medical record numbers, social security numbers, account numbers, any unique identifying number or code |
| FECHA | Dates | Dates |
| INST | Hospitals, healthcare centers or other institutions that might point to someone's location | - |

Header sections were included as a NE to ensure that they were not removed from the final text. These headers are necessary for further analysis, being key to extract the most relevant information of a radiology report.

### Corpus construction

The de-identification corpus consists of brain imaging radiology reports randomly extracted from the Medical Imaging Databank of the Valencian Region (BIMCV) database (19, 20), distributed among 17 health departments of the Valencian Region (Figure 1). A total of 7848 records were initially retrieved and automatically pre-annotated using the Spanish National Statistics Institute name and surname database (21), including those names with a frequency higher or equal to 20 in Spain, and a list of the hospital names in the Valencian Region. To ensure the presence of personal information in our corpus, a subset of reports with at least two ‘NAME’ tags was extracted. One-third of those reports were randomly selected to be manually corrected and annotated, with a final corpus of 692 records. The annotations were manually reviewed by three annotators, including finally all the NE tags.

**Fig. 1.**
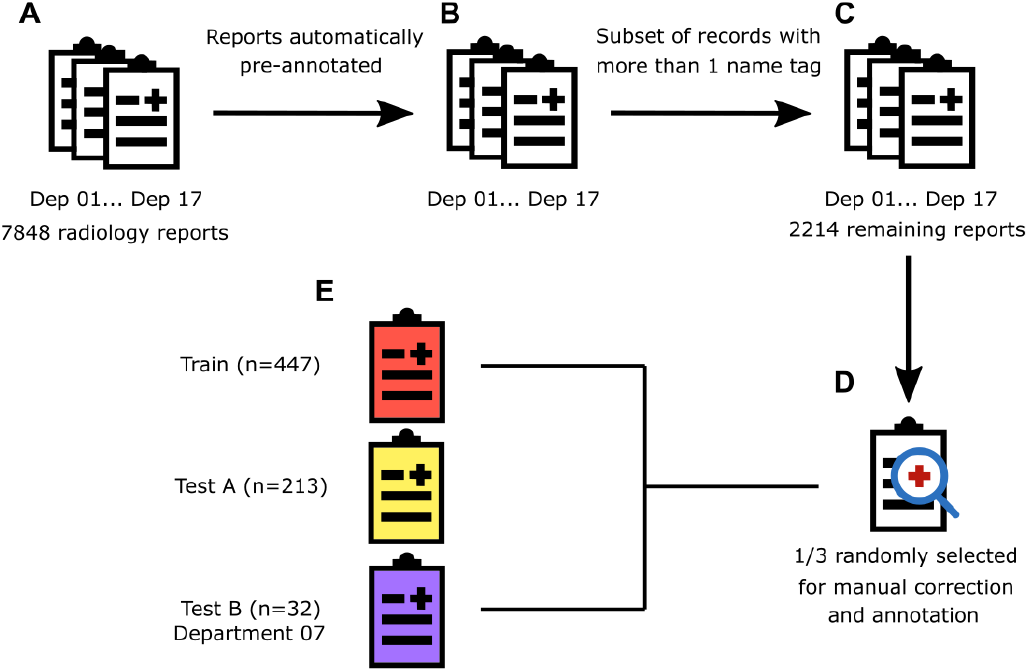
Data curation process and corpus preparation workflow. (A) 7848 radiology reports in total were retrieved from BIMCV database. (B) We used a custom Python script to automatically annotate the names, surnames and hospital names from radiology reports. (C) A subset of records was made meeting the condition that more than one ‘name’ tag was present, remaining 2214 reports. (D) Another subsetting was performed to randomly select one-third of reports to be manually annotated and corrected by three annotators. After the manual revision, 692 reports remain.(E) Ground Truth dataset was divided into 3 subsets: the training set included 447 reports, test A 213, and test B 32 reports from healthcare department number 7.

Radiology reports were not preprocessed so that they remain unchanged after the de-identification, apart from the identifying information. Although our radiology reports were mostly free-text sections preceded by headers, the 7th health department lacked headers and had an increased number of entities entirely out of context: this is, a name or a surname with no more text in an independent line, as shown in Figure 2. With this in mind, we divided our dataset into three sets, one for training and two for testing:

**Fig. 2.**
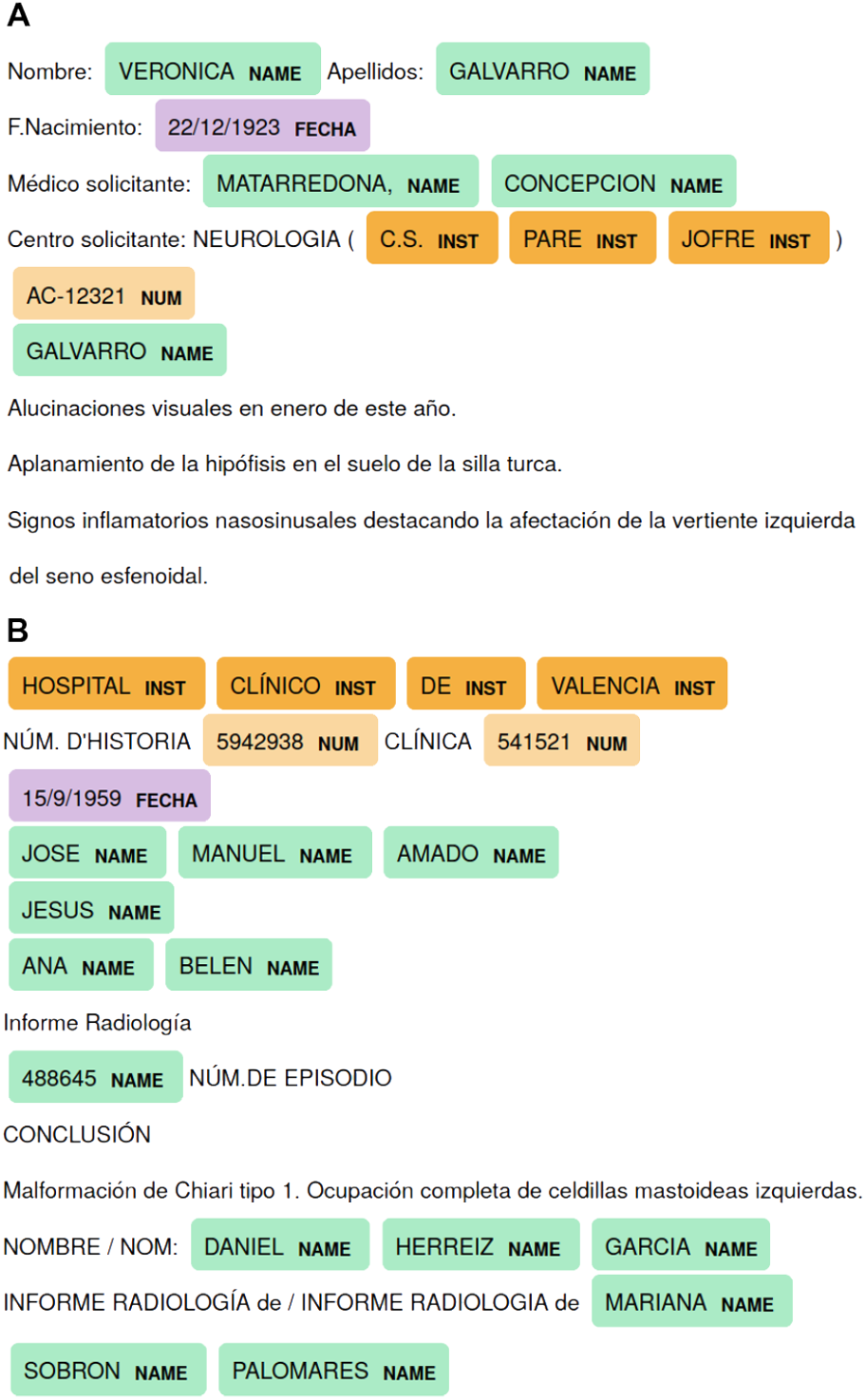
Partial examples of radiological records from Test A (A) and B (B). Test A has metadata headers clearly defined. In turn, test B has metadata headers in Valencian language and metadata information detached from these headers by a line break. Both structures include identifiable information in new lines without metadata headers.

- Training set, including 447 randomly selected records from all the departments.
- Test A set, including 213 randomly selected records from all the departments except 7th department.
- Test B set, including 32 randomly selected records from the 7th department.

Whereas both training and test A set present similar distribution of NEs (Table 2), test B shows an increase of addresses, locations and institutions. Having a separate test for department 7 allows us to check the performance of our method with highly unstructured data, with a distribution of NEs different from the training. As shown in Table 2, addresses and locations are the NEs with the lowest sample size.

**Table 2.** Number and percentage of annotations per corpus subset: Training, Test A and Test B.

|  | Training (words / %) | Test A (words / %) | Test B (words / %) |
| --- | --- | --- | --- |
| CAB | 1987 / 21.37% | 993 / 20.87% | 120 / 9.4% |
| NAME | 3286 / 35.34% | 1591 / 33.45% | 386 / 30.25% |
| DIR | 128 / 1.38% | 106 / 2.23% | 72 / 5.64% |
| LOC | 79 / 0.85% | 46 / 0.97% | 26 / 2.04% |
| NUM | 1159 / 12.47% | 585 / 12.29% | 143 / 11.21% |
| FECHA | 1655 / 17.79% | 897 / 18.86% | 300 / 23.51% |
| INST | 1004 / 10.80% | 539 / 11.33% | 229 / 17.95% |

To assess the performance of our final model with external data, we decided to incorporate 100 randomly selected clinical records from the MEDDOCAN task (16). These records have a different structure and are not related to radiology.

### NE randomization

We developed a methodology to randomize the PHIs found in a text, and applied it to the manually labelled dataset, obtaining a synthetic corpus. This methodology applies a set of rules depending on the NE associated with each tagged word. It is based on the substitution of tagged entities with new words randomly extracted from different databases available online:

- Spanish National Statistics Institute name and surname database (21), weighted by frequency
- Spanish National Statistics Institute municipal register database (22), weighted by population in 2019
- National Hospital Index (23)
- National Outpatients Clinic Index (24)
- Municipality addresses (25)

With the aim of avoiding the leakage of sensitive personal data, this methodology also checks that the randomly chosen word or number is not the same as the original one.

### Networks

A variety of neural networks were tested and evaluated, all of them designed for NER tasks. Three network architectures were based on Bidirectional Long Short-Term Memory (BiLSTM) layers, obtained from Guillaume Genthial’s GitHub repository (26):

- LSTM-CRF: GloVe vectors, BiLSTM and Conditional Random Fields (CRF) based on the work of Huang et al (27).
- LSTM-LSTM-CRF: GloVe vectors, character embeddings, BiLSTM for character embeddings, BiLSTM and CRF, based on the work of Lample et al (28).
- Conv-LSTM-CRF: GloVe vectors, character embeddings with 1D convolution and max pooling, BiLSTM and CRF, based on the work of Ma and Hovy (29).

These networks were trained with and without Exponential Moving Average (EMA) of the weights. We also trained a spaCy (30) NER model, based partly on the work of Lample et al (28) with dense embeddings along with Convolutional Neural Networks (CNNs) with an attention mechanism.

### Evaluation metrics

To assess the performance of the different models trained we computed precision, recall and F1-score metrics. These metrics can be defined as:

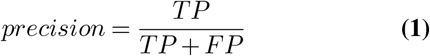

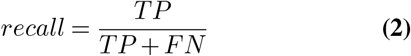

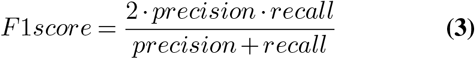

being TP the number of true positives, FP the number of false positives, and FN the number of false negatives.

To compute the amount of de-identification achieved by the model, we did not only applied these metrics to each NE, but to the set of words that should have been labelled as an identifying NE. With this approach, we obtained quantitative indicators of global de-identification.

## Results

First, models for each neural network were trained and then evaluated. Table 3 shows the mean global results of the different networks, given three replicates for each one.

**Table 3.** Evaluation metrics for the best model obtained with each of the different neural networks tested. Bold font highlights the best metric in each data subset.

| Model | Training |  |  | Test A |  |  | Test B |  |  |
| --- | --- | --- | --- | --- | --- | --- | --- | --- | --- |
|  | Precision | Recall | F1 | Precision | Recall | F1 | Precision | Recall | F1 |
| LSTM-CRF | 90.39 | 81.93 | 85.95 | 87.09 | 77.11 | 81.79 | 81.35 | 61.37 | 69.96 |
| LSTM-CRF with EMA | 91.19 | 84.15 | 87.53 | 87.05 | 78.49 | 82.55 | 71.48 | 59.65 | 64.96 |
| LSTM-LSTM-CRF | 99.20 | 98.79 | 98.99 | **98.13** | 97.18 | 97.66 | 93.01 | 90.94 | 91.96 |
| LSTM-LSTM-CRF with EMA | 99.06 | 98.96 | 99.01 | 98.00 | 97.34 | 97.67 | 94.20 | **91.10** | **92.63** |
| Conv-LSTM-CRF | 99.31 | 99.05 | 99.18 | 98.11 | 97.29 | 97.70 | **94.49** | 90.43 | 92.41 |
| Conv-LSTM-CRF with EMA | 99.17 | 99.05 | 99.11 | 98.08 | **97.36** | **97.72** | 93.72 | 90.64 | 92.15 |
| Spacy | **99.87** | **99.28** | **99.58** | 98.06 | 96.10 | 97.07 | 93.23 | 89.39 | 91.31 |

The recall is one of the most relevant evaluation metrics in any de-identification process (5), to avoid the leakage of sensitive information. Taking this into account, LSTM-LSTM-CRF with EMA shows the highest recall in test B, and Conv-LSTM-CRF with EMA in test A. Although these are the two best-performing networks in both test sets, we decided to include also spaCy for further analysis and leave outside the worst-performing architecture: LSTM-CRF.

The performance stats of each NE for LSTM-LSTM-CRF with EMA, Conv-LSTM-CRF with EMA and spaCy are displayed in tables 4, 5 and 6. Whereas in training set spaCy outperforms the other networks in every NE, in test A and test B results are more contested. Evaluating F1-score in test A, LSTM-LSTM-CRF classifies better dates, locations, names and numbers, while spaCy stands out with institutions. On the other hand, Conv-LSTM-CRF performs better with addresses and shows higher recall in names than LSTM-LSTM-CRF. In test B, the spaCy model shows better results in dates and better recall in institutions whereas LSTM-LSTM-CRF has a higher F1-score in institutions, locations and names. Conv-LSTM-CRF again performs better with addresses, but also with numbers and shows the highest recall in locations and names. When applying the models to MEDDOCAN dataset there’s a decay of the performance, although spaCy has higher recall rates in addresses, dates, institutions and name, whilst Conv-LSTM-CRF outperforms in locations and numbers.

**Table 4.** Evaluation metrics obtained with LSTM-LSTM-CRF with EMA model for each named entity.

|  | Training |  |  | Test A |  |  | Test B |  |  | MEDDOCAN |  |  |
| --- | --- | --- | --- | --- | --- | --- | --- | --- | --- | --- | --- | --- |
|  | Precision | Recall | F1 | Precision | Recall | F1 | Precision | Recall | F1 | Precision | Recall | F1 |
| CAB | 98.29 | 97.94 | 98.11 | 96.03 | 94.92 | 95.47 | 83.69 | 75.76 | 79.53 | 13.33 | 33.33 | 19.05 |
| DIR | 100 | 100 | 100 | 93.49 | 95.00 | 94.22 | 90.91 | 90.91 | 90.91 | 0.00 | 0.00 | 0.00 |
| FECHA | 99.74 | 99.64 | 99.69 | 98.93 | 99.20 | 99.07 | 96.65 | 94.83 | 95.74 | 74.95 | 86.34 | 80.20 |
| INST | 98.96 | 98.96 | 98.96 | 95.73 | 95.72 | 95.73 | 96.08 | 96.08 | 96.08 | 11.11 | 0.67 | 1.26 |
| LOC | 100 | 89.45 | 94.42 | 94.35 | 87.88 | 90.99 | 92.58 | 55.55 | 69.41 | 0.00 | 0.00 | 0.00 |
| NAME | 98.99 | 99.15 | 99.07 | 98.97 | 98.24 | 98.60 | 94.78 | 95.13 | 94.95 | 61.62 | 77.39 | 68.59 |
| NUM | 99.39 | 99.91 | 99.65 | 99.34 | 98.69 | 99.01 | 96.65 | 97.66 | 97.15 | 56.93 | 68.28 | 62.05 |
|  | 99.05 | 98.96 | 99.01 | 98.00 | 97.34 | 97.67 | 94.20 | 91.10 | 92.62 | 62.35 | 56.11 | 59.07 |

**Table 5.** Evaluation metrics obtained with Conv-LSTM-CRF with EMA model for each named entity.

|  | Training |  |  | Test A |  |  | Test B |  |  | MEDDOCAN |  |  |
| --- | --- | --- | --- | --- | --- | --- | --- | --- | --- | --- | --- | --- |
|  | Precision | Recall | F1 | Precision | Recall | F1 | Precision | Recall | F1 | Precision | Recall | F1 |
| CAB | 98.57 | 97.99 | 98.28 | 96.82 | 94.97 | 95.89 | 91.45 | 76.89 | 83.54 | 0.78 | 16.67 | 1.48 |
| DIR | 100 | 100 | 100 | 98.33 | 95.00 | 96.63 | 93.94 | 93.94 | 93.94 | 10.71 | 1.18 | 2.11 |
| FECHA | 99.71 | 99.78 | 99.74 | 98.79 | 99.13 | 98.96 | 95.77 | 93.21 | 94.47 | 82.28 | 86.21 | 84.18 |
| INST | 98.96 | 98.96 | 98.96 | 96.35 | 95.94 | 96.14 | 92.91 | 94.12 | 93.51 | 0.00 | 0.00 | 0.00 |
| LOC | 100 | 91.56 | 95.59 | 92.89 | 87.88 | 90.29 | 89.44 | 55.56 | 68.50 | 20.83 | 0.58 | 1.12 |
| NAME | 99.17 | 99.28 | 99.23 | 98.69 | 98.28 | 98.49 | 92.31 | 96.26 | 94.23 | 70.17 | 77.39 | 73.56 |
| NUM | 99.35 | 99.88 | 99.62 | 98.98 | 98.63 | 98.80 | 95.59 | 95.57 | 95.58 | 64.53 | 78.29 | 70.69 |
|  | 99.17 | 99.06 | 99.11 | 98.08 | 97.36 | 97.72 | 93.72 | 90.64 | 92.16 | 67.07 | 58.90 | 62.71 |

**Table 6.** Evaluation metrics obtained with spaCy model for each named entity.

|  | Training |  |  | Test A |  |  | Test B |  |  | MEDDOCAN |  |  |
| --- | --- | --- | --- | --- | --- | --- | --- | --- | --- | --- | --- | --- |
|  | Precision | Recall | F1 | Precision | Recall | F1 | Precision | Recall | F1 | Precision | Recall | F1 |
| CAB | 99.43 | 96.54 | 97.96 | 98.28 | 93.98 | 96.08 | 92.54 | 74.49 | 82.52 | 4.76 | 33.33 | 8.33 |
| DIR | 100 | 100 | 100 | 94.28 | 63.96 | 76.01 | 87.79 | 74.77 | 61.46 | 43.15 | 4.47 | 8.01 |
| FECHA | 100 | 100 | 100 | 98.54 | 99.04 | 98.78 | 98.20 | 97.53 | 97.86 | 51.39 | 89.41 | 65.13 |
| INST | 99.97 | 99.96 | 99.98 | 98.19 | 97.24 | 97.71 | 93.50 | 98.00 | 95.69 | 45.72 | 12.28 | 19.27 |
| LOC | 100 | 100 | 100 | 76.64 | 54.66 | 63.80 | 61.04 | 26.85 | 36.79 | 7.19 | 0.32 | 0.59 |
| NAME | 100 | 99.99 | 99.99 | 98.34 | 98.28 | 98.31 | 88.78 | 94.29 | 93.19 | 75.62 | 83.91 | 79.23 |
| NUM | 100 | 100 | 100 | 97.81 | 95.65 | 96.72 | 95.11 | 87.56 | 91.18 | 68.50 | 60.32 | 63.99 |
|  | 99.87 | 99.28 | 99.58 | 98.06 | 96.10 | 97.08 | 93.23 | 89.39 | 91.31 | 65.63 | 55.37 | 59.98 |

Given that our aim was not to correctly classify NE, but to completely remove sensitive information from the text, global de-identification metrics were computed (Table 7). Conv-LSTM-CRF with EMA shows better recall in test A and test B sets (Figure 3), whilst LSTM-LSTM-CRF has higher F1-score on test B. On MEDDOCAN data the model that better maintains recall and F1-score is LSTM-LSTM-CRF (Figure 3, Table 7). To assess the performance of our models with external data, we wanted to apply the models generated at MEDDOCAN to our data. Only one of the participants made their models available (31), being one of the implemented networks spaCy. Their spaCy model achieved a precision of 87.89% and 80.31%, a recall of 42.66% and 26.54%, and an F1-score of 57.44% and 39.89% in our test A and our test B, respectively (Table 7).

**Table 7.** Global de-identification metrics for LSTM-LSTM-CRF, Conv-LSTM-CRF, spaCy and the model retrieved from MEDDOCAN. (*) : Results extracted from the original publication (31).

|  | Test A |  |  | Test B |  |  | MEDDOCAN |  |  |
| --- | --- | --- | --- | --- | --- | --- | --- | --- | --- |
|  | Precision | Recall | F1 | Precision | Recall | F1 | Precision | Recall | F1 |
| LSTM-LSTM with EMA | 99.66 | 99.29 | 99.48 | 99.08 | 97.18 | 98.10 | 98.09 | 69.18 | 81.13 |
| Conv-LSTM-CRF with EMA | 99.58 | 99.42 | 99.50 | 98.18 | 97.43 | 97.80 | 97.11 | 67.10 | 79.36 |
| spaCy | 99.28 | 94.15 | 96.64 | 95.69 | 91.18 | 93.38 | 84.96 | 61.69 | 71.48 |
| MEDDOCAN model | 87.89 | 42.66 | 57.44 | 80.31 | 26.54 | 38.89 | 96.70* | 95.30* | 96.60* |

**Fig. 3.**
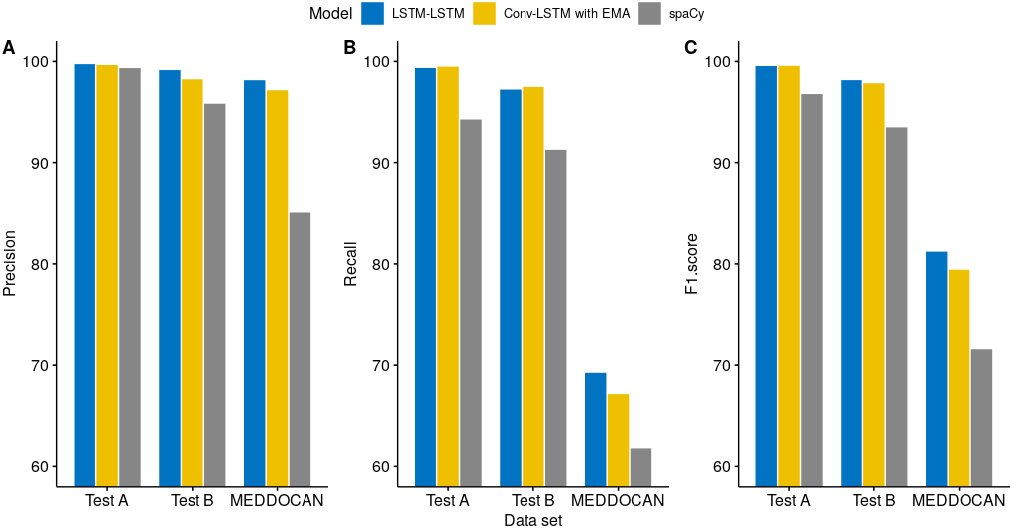
Precision (A), recall (B) and F1-score (C) for the three best performing architectures, LSTM-LSTM-CRF with EMA (blue), Conv-LSTM-CRF with EMA (yellow) and spaCy (grey) by data subset.

## Discussion

This work has defined and evaluated a methodology based on NER to de-identify radiology reports in Spanish language. In comparison with traditional approaches based on regular expressions, NLP and neural networks do not underperform due to human misspellings or the absence of a clear and repeated structure. Neural networks are also context-dependent, and words like Parkinson will be detected as a ‘NAME’ entity when used as a surname but left unchanged when used as a medical word, avoiding the loss of meaningful information. The main drawback of this methodology is the requirement of a learning corpus of de-identified reports, which is not necessary for regular expression-based strategies. Although the curation of a corpus is a tedious and methodical task, there is no need for a big dataset: with a training set of 447 texts, we achieved a suitable performance.

Neural networks should be trained with a corpus diverse in structure to avoid overfitting. Machine learning models tend to learn the structure or format of the text, finding the position of words containing sensible data when performing deidentification. If a model was trained with a corpus with a determined structure, it will only be able to de-identify similarly-formatted texts. By comparing our spaCy model with the spaCy model retrieved from MEDDOCAN (31), we show the high impact that text structure has in the outcome. The MEDDOCAN training set was similar in size to ours (500 and 447 texts with a median of 20 and 22 lines per text, respectively), but their text structure was highly defined and invariant. With a training set diverse in its structure we can obtain higher recall and precision in external data, generating a de-identification model better prepared to deal with new data. Figure 2 illustrates the structure and format diversity of radiological reports between health departments included in our dataset.

Considering that the recall metric assesses the capability to avoid the leakage of sensitive information of a model, we propose LSTM-LSTM-CRF with EMA as the best neural network to address a de-identification task based on NER. This neural network showed higher statistics in all three pro-posed test sets, and its recall in test A and test B are comparable to those obtained with Conv-LSTM-CRF with EMA. Thus, we expect LSTM-LSTM-CRF with EMA to behave optimally when presenting new data to it. Although its recall is very good, it is not perfect. When new radiology reports from the Valencian Region are included in BIMCV database, 97.18% of recall in test B means that almost 3% of identifying words will remain in the text. It might not be enough to re-identify the patient: could be left only a surname, a city name, or a part of an address. To ensure that the identity of a patient is not restorable, a final check of the texts by an authorized person remains necessary. Nevertheless, we propose a randomization strategy to change the identified NEs for synthetic ones of the same category. This strategy masks the identifying words left by the neural network with synthetic information, making it more difficult to discern between real and synthetic identifying words than by simply erasing words (Figure 4). Further efforts need to be done to validate whether this strategy makes original information unretrievable or not.

**Fig. 4.**
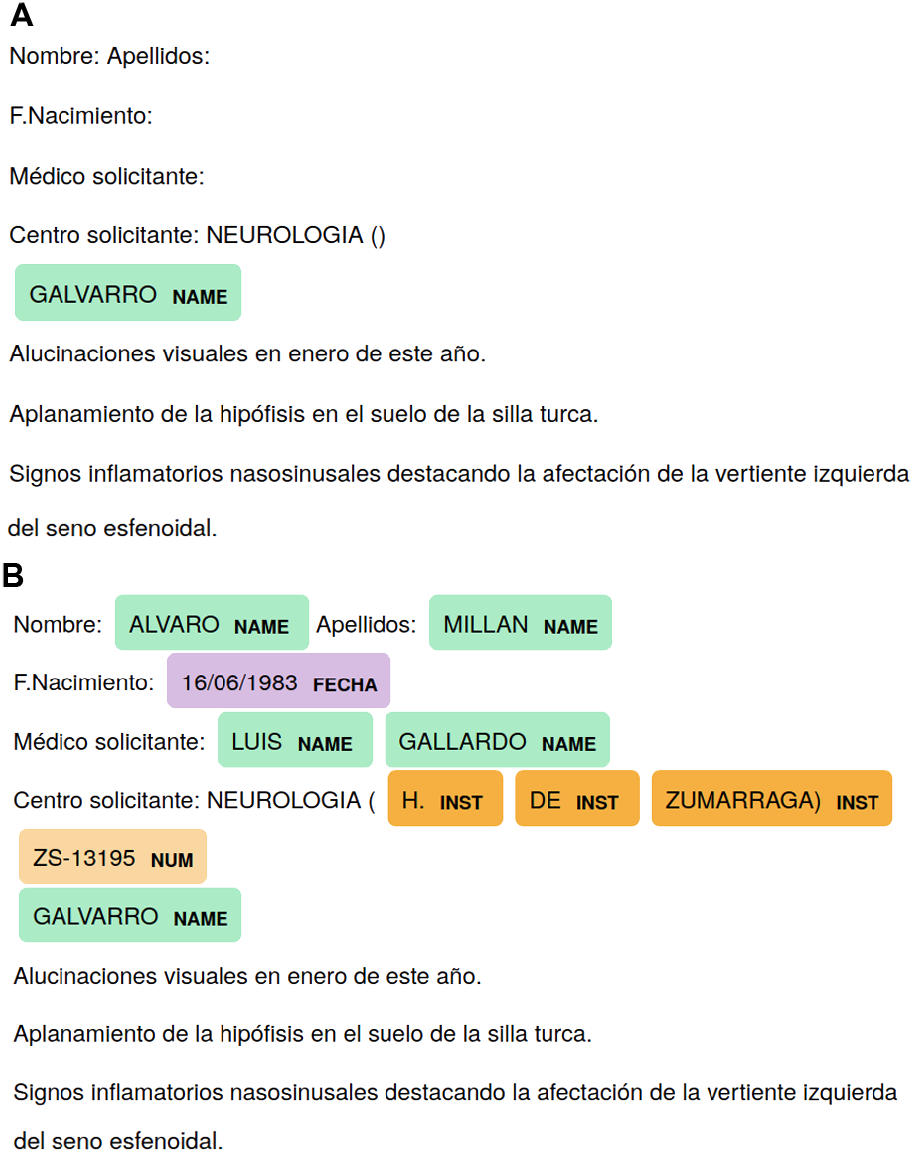
Anonymization strategies. When applying word elimination (A) errors are easily detectable whereas with synthetic substitution (B) any mistake is hidden with randomized synthetic information.

## Conclusions

Medical texts hold great potential for research, but legal and privacy concerns arise with its use, even more, when institutions external to the hospital are involved. Realworld medical texts tend to be semi-structured with free text that includes sensible information, thus classical deidentification approaches based on regular expressions are not good enough. We propose a robust and flexible methodology based on NER for Spanish medical texts, tested on radiology reports from the Valencian Region. This method is generic and relatively simple and can be easily generalizable to other Spanish medical texts by re-training the network with additional data. We believe it can be also replicated in other languages, at least Romance derived languages, being the easiest network to implement spaCy, although it is not the best performing. The proposed de-identification methodology still missed identifiers after training, thus a final check of the texts by an authorized person remains necessary. Nevertheless, we believe a combination of NER with the generation of synthetic data will make it virtually impossible to extract real identifying words from the text. Further efforts need to be done to assess and test this hypothesis.

## Data Availability

The data that support the findings of this study are available from BIMCV but restrictions apply to the availability of these data under a research use agreement. Data access can be requested at http://bimcv.cipf.es/.
Supplementary information and code are available online at https://github.com/BIMCV-CSUSP/DiSMed.

https://bimcv.cipf.es/bimcv-projects/dismed/

## ACKNOWLEDGEMENTS

This article describes work undertaken in the context of the DeepHealth project, “Deep-Learning and HPC to Boost Biomedical Applications for Health” (https://deephealth-project.eu/) which has received funding from the European Union’s Horizon 2020 research and innovation programme under grant agreement No 825111”. The contents of this publication reflect only the author’s view, can in no way be taken to reflect the views of the European Union and the Community is not liable for any use that may be made of the information contained therein.

## ETHICS APPROVAL

The study was approved by the local institutional ethics committee DGSP-CSISP NÚM. 20190503/12.

## AVAILABILITY OF DATA AND MATERIALS

The data that support the findings of this study are available from BIMCV but restrictions apply to the availability of these data under a research use agreement. Data access can be requested at http://bimcv.cipf.es/.

Supplementary information and code are available online in GitHub.

- Project name: DiSMed - De-identifying Spanish medical texts
- Project home page: https://github.com/BIMCV-CSUSP/DiSMed
- Operating system(s): Platform independent
- Programming language: Python
- Other requirements: Python (version≥ 3.5). DiSMed imports the following Python non-built-in libraries: pandas, numpy, codecs, spacy, tensorflow (version *<* 2)
- License: MIT

